# Study Protocol: Risk and resilience factors in problematic internet use among Sámi and non-Sámi adolescents in Finnmark, Arctic Norway: the role of social norms and ethnic identity

**DOI:** 10.64898/2026.08.12.26360242

**Authors:** Shiho Hansen, Snefrid Møllersen, Anna Rita Spein, Anne Cecilie Jávo

**Affiliations:** Sámi Norwegian National Advisory Unit for Mental Health and Substance Use, Finnmark Hospital Trust, Karasjok, Norway; Centre for Sami Health Research, UiT Arctic University of Norway, Tromsø Norway

## Abstract

Problematic Internet Use (PIU)—marked by compulsive or maladaptive online behavior—is an emerging public health issue among adolescents and is associated with psychological distress, social difficulties, and academic problems. In Finnmark County, Norway’s northernmost and ethnically diverse region, limited research has examined the underlying mechanisms of PIU among Sámi and non-Sámi youth, despite increasing levels of digital engagement. This study protocol outlines a population-based cross-sectional survey investigating the associations between social norms (descriptive and injunctive), ethnic identity, and ethnicity-based discrimination in relation to PIU among Sámi and non-Sámi adolescents in Finnmark. Guided by Social Norm Theory and Ethnic Identity Theory, the study aims to examine risk and resilience factors associated with adolescents’ digital behavior in a geographically sparsely populated, multiethnic region. A population-based, cross-sectional school survey will include all upper secondary school students in Finnmark County (N ≈ 2,230). A culturally adapted, bilingual questionnaire (Northern Sámi – Norwegian) will measure problematic internet use, perceived social norms in family, peer, and school contexts, ethnic identity, ethnicity-based discrimination, positive internet use, and key covariates. Ethnicity will be classified based on indicators of Sámi language use and self-identification. Data will be prepared using prespecified quality procedures and analyzed with partial least squares structural equation modeling (PLS-SEM) to examine associations between social norms, ethnic identity, ethnicity-based discrimination, and internet use outcomes, including mediation and moderation. Group differences between Sámi and non-Sámi adolescents will be assessed using PLS Multi-Group Analysis. The findings may inform the development of culturally appropriate approaches to screening, prevention, and early intervention, and are relevant for mental health services, school-based programs, and public health strategies targeting Indigenous youth in rural and semi-rural regions.

## Introduction

### Adolescents’ problematic internet use

Over the past decade, social media and digital platforms have become central to adolescents’ lives worldwide [1]. While media use offers benefits—such as early learning, access to knowledge [2], peer connections [3], and exposure to diverse perspectives [4] —it also poses risks, including addiction, mental and physical health challenges, sleep disruption, and exposure to harmful content [2, 5].

A particularly concerning consequence of excessive internet use is Problematic Internet Use (PIU)— understood in this protocol as a pattern of excessive, compulsive, and difficult-to-control online behavior—is an increasing public health concern [6] and a growing challenge in adolescent mental health care [7]. PIU may occur across a range of online contexts, including gaming, gambling, pornography consumption, and cyberbullying, and is associated with psychological distress; it commonly co-occurs with affective symptoms such as depression and anxiety [5], and shares neurobiological correlates with addictive and impulse-control disorders [8]. PIU is further associated with higher levels of engagement in risky behaviors (e.g., cyberbullying and desensitization to aggression through violent media exposure) [9], sleep disturbances [10], and impairments in social and school functioning [6], including reduced face-to-face interaction [11]. Despite these risks, clinical and research efforts are constrained by a lack of consensus on diagnostic criteria and the absence of culturally adapted, empirically grounded guidelines for distinguishing clinically concerning from normative internet use among youth [7].

Environmental factors such as family, peers, and schools significantly shape adolescents’ digital behaviors [6]. Family dynamics, including parent-child relationships and media habits, play a crucial role [12], with close relationships and open communication reducing PIU [13]. Peers also significantly shape adolescents’ online behaviors. Peer communication pressure impacts social media use, contributing to clustering effects and fostering internet and gaming addictions [13, 14]. Friendship quality and social support show differing effects, with strong friendships enhancing internet use and poor support linked to PIU [15]. Schools also moderate PIU through supportive environments [16]. For example, shared gaming interests within classrooms and during leisure time may foster social connection and reduce PIU symptoms [17].

### Internet use in Finnmark

Challenges related to PIU are particularly relevant in Finnmark, Norway’s northernmost county. Recent quantitative findings indicate that adolescents in this region spend more time on social media and online gaming than the national average, with a marked increase over the past decade [18]. Digital media use in Finnmark is shaped by environmental factors such as family dynamics, school connectedness, and satisfaction with the local community area. A qualitative study has shown strong place attachment among Indigenous Sami secondary and high school students [19]. Social media use, in particular, is associated with peer influence, emotional health, adverse experiences, and depressive symptoms [20].

Despite these patterns, existing research has largely focused on usage frequency and duration, with limited attention to problematic behaviors. More broadly, ethnicity and geographical context have received limited consideration, despite the region’s multiethnic composition, which includes the Indigenous Sámi population [21]. Ethnic differences in problematic internet use and its associated factors remain underexplored. These gaps underscore the need for closer examination of how digital media use is associated with outcomes among adolescents across different ethnic backgrounds.

### Internet use and Indigenous adolescents

Studies on Indigenous adolescents globally highlight both protective and risk factors associated with internet use. In Australia, research has shown that social media and digital technologies play a dual role in the lives of Aboriginal youth, fostering identity expression, empowerment, and community connection, while also exposing them to ethnicity-related cyberbullying and harmful content [22].

Among Inuit youth in Canada, social networking platforms, particularly Facebook, are essential for identity affirmation, sociocultural dialogue, and the preservation of traditions, suggesting that digital media has become integral to cultural identity rather than merely an external influence [23]. For Indigenous adolescents attending boarding schools in remote areas of Australia, mobile phones are widely used for communication and cultural connection, whereas television and video games tend to play a less significant role [24]. Cyberbullying remains a significant concern among Indigenous adolescents. In Canada, cyberbullying victimization has been strongly linked to increased anxiety and stress among Indigenous youth [25]. Similarly, a study of First Nations youth across Canada highlights the psychological distress associated with racial online victimization, particularly among females [26]. A large U.S. study of college students (N = 417,780) found that the prevalence of PIU was higher among Non-Hispanic American Indian/Alaskan Native/Native Hawaiian students than among Non-Hispanic White students [27].

Sámi youth in Norway similarly face exclusion and prejudice in both social and institutional settings, which has been linked to heightened psychological distress [28]. Mental health of Sámi adolescents is affected by ethnicity-based discrimination and bullying [29–31], indicating increased vulnerability to negative experiences in peer and social environments, including online contexts. Taken together with findings from other Indigenous populations, these patterns suggest that online spaces may function both as arenas for cultural affirmation, social connection, and support, but also as contexts where stress and social pressure occur.

### Social norms

Behaviors often cluster within personal relationships and social contexts, with social norms playing a central role in shaping these patterns [32]. Social norms are commonly defined as “the rules and standards that are understood by members of a group and that guide and/or constrain social behavior without the force of laws” [33]. Alternatively, they are described as “a rule of behavior such that individuals prefer to conform to it on the condition that they believe (a) most people in their reference network conform to it and (b) most people in their reference network believe they ought to conform to it” [34]. Several key theories have contributed to our understanding of social norms, including the focus theory of normative conduct [33], the theory of social norms [32], and the theory of normative social behavior (TNSB) [35]. These theories distinguish two types of social norms: descriptive norms, which refer to beliefs about what others typically do in a given situation, and injunctive norms, which refer to belief about what behaviors other people approve or disapprove of in that context [33]. Both types of norms can significantly influence behavior and decision-making processes [32]. In particular, the TNSB provides a comprehensive framework for understanding how descriptive norms affect behavior, and how this influence is moderated by injunctive norms [35].

Social norms across contexts such as family, peers, and schools influence adolescent risk-taking behavior [36]. For instance, research on adolescent alcohol consumption shows that descriptive norms are strong predictors of drinking behavior [37]. School-wide descriptive norms have been linked to increased substance use over time, while injunctive norms tend to influence behavior indirectly [38].

Although research on social norms and PIU remains limited, existing studies on digital behaviors suggest similar trends. Descriptive norms have been identified as stronger predictors of risky sexual online behavior [39] and illegal smartphone use while driving [40]. Injunctive norms in these contexts generally moderate behavior by shaping attitudes and reinforcing patterns, rather than directly determining actions. Overall, the evidence suggests that while both types of norms are influential, descriptive norms consistently exert more direct effects on behavior, whereas injunctive norms function primarily as moderators, reinforcing attitudes and behavioral tendencies.

### Sámi adolescents and social norms in internet use

Social norms are shaped by individual differences and social interactions within reference groups, which can influence conforming to group behavior and individual decision-making [32, 33]. In Sámi communities, reference groups often extend beyond the nuclear family to include a broader kinship network. Extended family operates under a multi-parent principle [41], where not only parents but also ritual kin share child-rearing responsibilities. This dynamic family network provides emotional, practical, and cultural support [42, 43]. Sámi child-rearing practices prioritize independence, resilience, and indirect control, blending autonomy with collectivist values [44, 45]. Alongside family and kinship networks, school also functions as an important reference context for Sámi adolescents, shaped by both educational opportunities and stressors. Prior research indicates strong educational aspirations among some Sami groups, but lower overall school well-being, particularly in Sámi-dominated inland areas and those affiliated to reindeer herding [19, 46].

These cultural foundations shape both descriptive and injunctive norms around internet use. Traditional Sámi parenting emphasizes autonomy and indirect control, as adolescents are expected to self-regulate. This may reduce parental intervention, increasing the risk of persistent PIU. At the same time, cultural values can be protective, as the strong cultural emphasis on child self-determination and internal locus of control in Sámi child-rearing may foster resilience and responsible behaviors [44]. In such cases, injunctive norms reinforce descriptive norms, strengthening their influence on behavior through perceived social approval.

### Ethnic identity

Ethnic identity— an individual’s sense of being a person who is defined by membership in a specific ethnic group, typically defined by shared social, cultural, linguistic, and religious factors [47]—plays a crucial role in shaping behaviors [48, 49]. According to Phinney’s Ethnic Identity Theory [50], it is not fixed but evolves over time through an active process beginning in early childhood and continuing into adulthood. The theory conceptualizes ethnic identity in terms of key dimensions, primarily exploration and commitment. Exploration involves engaging in activities and experiences that increase understanding of one’s ethnic background, while commitment reflects a sense of belonging and emotional attachment to one’s ethnic group. Through ongoing exploration and commitment, individuals may develop a more clearly defined or achieved sense of ethnic identity [50].

### Ethnic identity among Sámi adolescents

In Northern Norway, stronger ethnic identity among Sámi adolescents has been associated with lower engagement in risk behaviors and better mental health outcomes. Lower cultural orientation— often linked to assimilation—has been associated with higher rates of smoking and alcohol use in late adolescence [51]. Sámi adolescents reporting stronger ethnic identity, reflecting higher levels of commitment and attachment, show better mental health, whereas identity insecurity is linked to increased psychological vulnerability [28]. Studies further indicate that cultural practices, extended family networks, and connection to land reinforce ethnic identity and well-being [43]. Ethnic commitment is particularly strong among Sámi youth, especially females, in Sámi-dominated inland areas, while weaker identity is observed among adolescents with multiethnic backgrounds and no affiliation with reindeer herding [52]. Findings from the North Norwegian Youth Study show that ethnic identity varies by context, with the strongest Sámi identity in Sámi-dominated inland areas and the strongest Norwegian identity in coastal areas [46]. Importantly, anxiety and depression were more closely associated with ethnic identity than with ethnic group itself [31].

### Ethnic identity and PIU

Although ethnic identity is well-established as a protective factor against various risk behaviors, its role in PIU among Sámi adolescents remains unexamined. However, research on other Arctic and Indigenous populations suggests that ethnic identity and ethnic group participation may help mitigate PIU. For example, Aboriginal youth in Australia use social media for identity affirmation and community building, though cyberbullying remains a concern [22]. Inuit youth in Canada engage with digital platforms for cultural expression and tradition preservation [23]. Among First Nations youth across Canada, participation in cultural events within their group buffered cyberbullying distress, whereas passive ethnic group appreciation did not [26]. These findings highlight the significance of ethnic identity in shaping digital behaviors.

### Theoretical framework

Drawing from the literature, this study applies an integrated theoretical framework combining Social Norm Theory and Ethnic Identity Theory to understand PIU among Sámi and non-Sámi adolescents in Finnmark.

Social Norm Theory [32, 33, 35] distinguishes between *descriptive norms* (what people commonly do) and *injunctive norms* (what people believe ought to be done). This framework provides insight into how adolescents’ online behaviors are influenced by perceived norms across family, peer, and school contexts, and how injunctive norms may moderate the effects of descriptive norms.

Ethnic Identity Theory, particularly Phinney’s model, conceptualizes ethnic identity as a set of key dimensions—primarily exploration and commitment—that shape how individuals understand and relate to their ethnic group [50]. Among minority youth, including Sámi adolescents, strong ethnic identity can function as a protective factor, while identity insecurity may increase vulnerability to various psychosocial risks. Together, these theories allow for an examination of how social norms and ethnic identity interact to influence PIU. The framework supports analysis of how normative pressures and ethnic identity jointly shape adolescents’ digital behaviors, acting as both risk and resilience factors.

### Purpose of the study

This study aims to investigate the interplay of risk and protective factors influencing problematic internet use (PIU) among Indigenous Sámi and non-Sámi adolescents in Finnmark, as depicted in Fig. Specifically, the study will:

1. Assess the prevalence of both PIU and beneficial internet use among Sámi and non-Sámi adolescents.
2. Examine the extent and association of descriptive and injunctive social norms across family, peer, and school contexts with PIU, explore the differences in these associations between Sámi and non-Sámi adolescents.
3. Investigate whether a strong ethnic identity among Sámi adolescents serves as a protective factor, while experiences of ethnicity-based discrimination contribute to PIU.
4. Integrate social norms, ethnic identity, and victimization to examine their direct, moderating, and mediating associations with problematic internet use (PIU), and to compare these pathways between Sámi and non-Sámi adolescents.

Ethnic community context is accounted for by adjusting for municipal differences within Finnmark.

**Fig 1.**
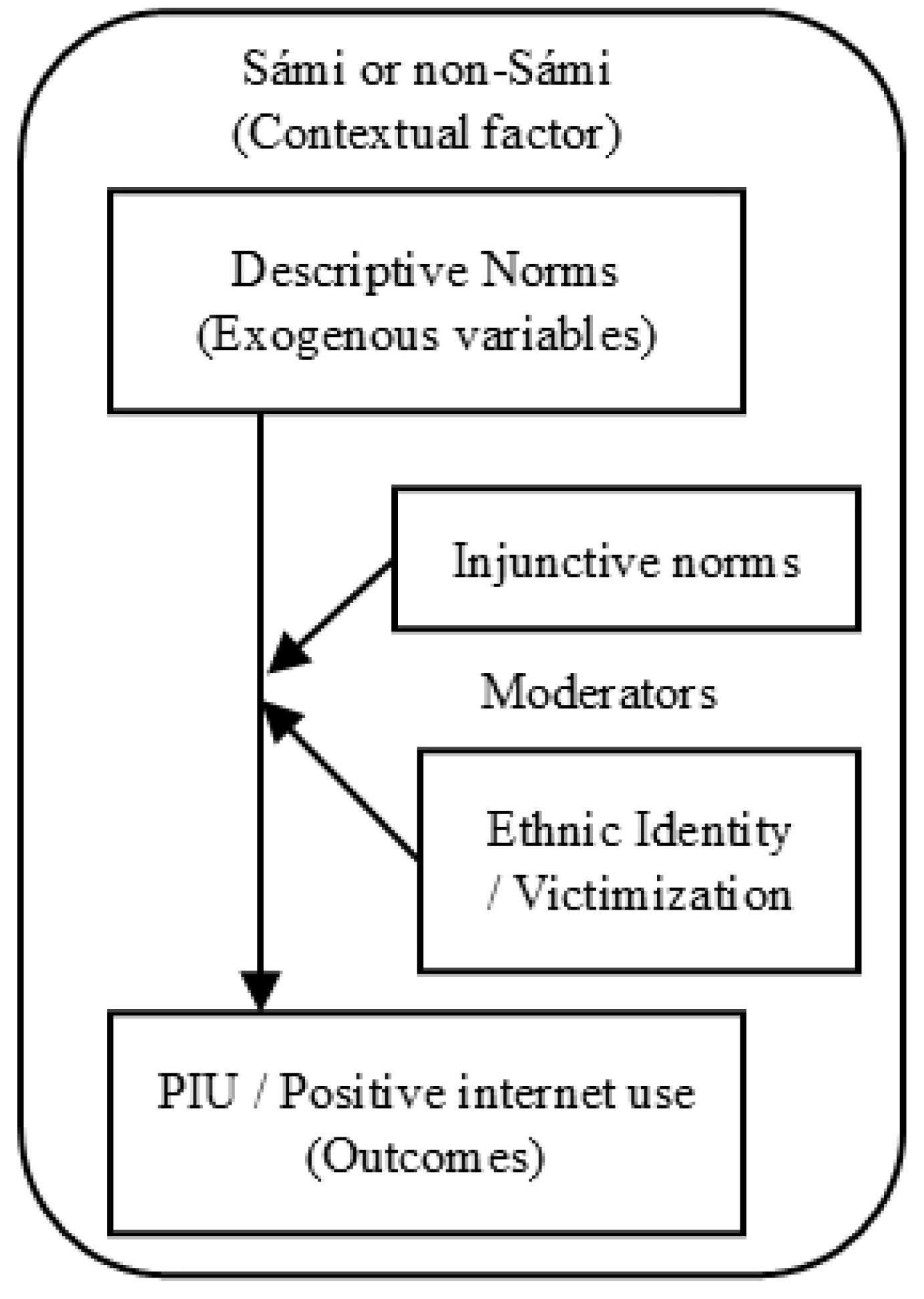
Conceptual framework of the study.

### Hypothesis

**H1**: Higher levels of descriptive norms of PIU within family, peer, and school environments will be associated with increased PIU. Injunctive norms will moderate this relationship. The strength of these associations will differ between Sámi and non-Sámi adolescents.

**H2**: Among Sámi adolescents, strong ethnic identity will serve as a protective factor by moderating the relationship between social norms, victimization, and PIU. Specifically, stronger ethnic identity will be associated with lower levels of PIU and will buffer the impact of ethnicity-based discrimination.

**H3:** Sámi and non-Sámi adolescents who report stronger ethnic identity and supportive social norms will also report higher levels of positive internet use, such as using the internet to maintain social connections, seek social support and emotional connection, and engage with ethnic identity.

## Material and methods

### Research design

This study employs a cross-sectional design with a quantitative approach and adopts an interdisciplinary perspective by integrating insights from psychology, psychiatry, sociology, Indigenous studies, and public health. The theoretical framework reflects this integration by combining Social Norm Theory from behavioral science with Ethnic Identity Theory from cultural and developmental psychology.

### Participants and sampling methods

The participants will be high school students (VG1–VG3, equivalent to 11th–13th grade) in Finnmark County. According to the Norwegian Directorate for Education and Training (*Utdanningsdirektoratet*), the total number of enrolled high school students in Finnmark County for the 2025–2026 school year was 2 320 [53]. Based on response rates (60–70%) observed in the National Norwegian Youth Survey (*Ungdata*)—a widely used national monitoring system for adolescent health and well-being [54] — we estimate that approximately 1,400–1,500 students will participate.

Because no official statistics on ethnic identity are available at the individual level, precise estimates of the Sámi student population in Finnmark cannot be derived. Feasibility is therefore assessed using official enrollment figures for upper secondary schools, which directly reflect the study’s sampling frame. High schools located in municipalities covered by both the Sámi Language Act and the Sámi Parliament’s administrative area are commonly associated with a higher proportion of Sámi residents. These municipalities include Kautokeino, Karasjok, Lakselv (Porsanger), and Tana.

Together, these schools—along with Sámi adolescents living outside these areas—are estimated to include more students than the minimum sample size required for PLS SEM analyses (see *Statistical approach and sample size calculation*).

Given the relatively small and geographically defined population, the absence of a reliable ethnic sampling frame, and the feasibility of recruiting students through participating schools, the study applies a total population (census based) recruitment strategy including all high schools in Finnmark County (VG1–VG3; equivalent to 11th–13th). This approach minimizes selection bias and ensures sufficient statistical power for the planned analyses, including comparisons between Sámi and non-Sámi adolescents.

### Statistical approach and sample size calculation

This study will utilize Partial Least Squares Structural Equation Modeling (PLS-SEM) to ensure adequate statistical power and robustness. PLS-SEM is well-suited for analyzing complex models involving latent constructs. The method is suited for this study because it yields reliable results with smaller sample sizes, maintains high statistical power, does not require normally distributed data, and is resilient to measurement error. Additionally, PLS-SEM employs bootstrapping to estimate confidence intervals for the path coefficients, an assumption-free method that helps reduce bias in these estimations [55]. In studies grounded in social norms theory, mediation and moderation analyses are commonly used to examine the relationships between normative beliefs and behavioral outcomes across family, peer, and school contexts [35, 36, 39, 40]. PLS-SEM is a robust analytical approach for testing such relationships, making it particularly appropriate for this study’s theoretical and methodological framework.

The choice of PLS-SEM is aligned with the census-based design by maximizing statistical power while accommodating typical survey data distributions. First, estimating the structural model on the full population of interest (i.e., all eligible students) maximizes statistical power at the model level while accommodating the non-normal distributions typical of survey data. Second, planned comparisons between Sámi and non-Sámi adolescents will be conducted using PLS Multi-Group Analysis (PLS-MGA) with bootstrapping, which is robust to unequal group sizes and therefore well suited to the demographic composition of the study population.

To determine the minimum required sample size for PLS-SEM analysis, this study will apply the inverse square root method to provide precise estimates [56]. The inverse square root method is widely used due to its conservative nature and simplicity. Assuming 80% power, a 5% significance level, and path coefficients between 0.11 and 0.20, the estimated minimum required sample size is 155 participants [55, 56]. Given the anticipated overall sample size and the census-based recruitment strategy, this requirement is expected to be met for both Sámi and non-Sámi adolescents, supporting stable parameter estimation and planned between-group analyses.

Because an imbalance in group sizes between Sámi and non-Sámi students is expected due to demographic distribution, PLS-MGA with bootstrapping will be used for subgroup comparisons.

PLS-MGA is a non-parametric procedure that is robust to unequal group sizes and allows reliable comparison of group-specific structural path estimates [55, 57].

### Work packages

The project is structured into four main work packages (WP):

WP1: Instrument Development– Led by the core team in collaboration with high school students

WP2: Data Collection – Coordinated with participating schools and supported by research assistants

WP3: Data Analysis – Led by the core team with support from statistical experts

WP4: Dissemination – Includes academic publications, conference presentations, and local outreach.

### WP1: Instrument development

The study questionnaire has been developed through a structured, multi-phase process to ensure cultural sensitivity and linguistic appropriateness. The instrument combines validated tools and newly developed items, adapting established theories and prior survey research. Constructs such as PIU, social norms, and ethnic identity will be measured using Likert-type scales or a Visual Analog Scale— a type of psychometric scale that employs a continuous measurement indicator rather than multiple discrete options. Higher values will indicate more undesirable attitudes or behaviors. The questionnaire is available in both Norwegian and Northern Sámi language to enhance inclusivity, allowing Sámi-speaking students to participate in their native language. The development process consists of the following key steps:

### Cultural adaptation, translation, and pretesting (completed in the first quarter of 2026)

The questionnaire draws from validated Norwegian instruments, including those used in the North Norwegian Youth Study (NNYS) [46], a longitudinal epidemiological study conducted in the three northernmost counties of Norway between 1994–1995 and 1997–1998; the Norwegian Arctic Adolescent Health Study (NAAHS) [31], which was carried out in the same region beginning in 2003; and the National Norwegian Youth Survey, to assess ethnicity, ethnic identity, and experiences of discrimination. The PIU questionnaire has been culturally adapted for Sámi and non-Sámi students in Finnmark, while items measuring social norms were developed based on relevant theories and prior research. Input from group interviews with 15 Sámi and non-Sámi high school students informed the adaptation of PIU and social norms items, ensuring cultural relevance and comprehensibility. Items not previously translated into the Northern Sámi language—particularly those related to PIU and social norms—were translated from Norwegian using a forward–backward translation method to ensure linguistic accuracy and conceptual equivalence.

The draft questionnaire will undergo a pretest with approximately 30 Sámi and non-Sámi students to evaluate the clarity, comprehension, and usability of the items. The pretest aims to identify potential issues with question interpretation, the estimated completion time, and any participant burden. Feedback from participants will guide the refinement of wording, layout, and structure to improve the overall quality of the questionnaire.

### Pilot testing and finalization of the questionnaire

Following the pretest, a pilot test will be conducted with a separate group of approximately 30 Sámi and non-Sámi students. The goal of the pilot test is to assess the preliminary reliability and validity of the questionnaire. Internal consistency will be measured using Cronbach’s alpha, while exploratory factor analysis will be used to explore the construct structure of the instruments. The results from the pilot test, combined with participant feedback, will be used to make final revisions to the questionnaire.

Students participating in the pretest and pilot test will not be included in the main study sample (WP2: Data Collection), as these activities are conducted during the 2025–2026 school year primarily with VG3 students (equivalent to 13th grade), who will have graduated before the 2027 data collection begins. This ensures that there is no overlap between pilot participants and the total population sample.

Upon completion of the study and publication of results, the Norwegian and Northern Sámi versions of the questionnaire will be made openly accessible for research purposes.

### WP1.1: Measuring instruments

#### Classification of Sámi and non-Sámi adolescents

Ethnicity is a multidimensional construct that encompasses ancestry, language, cultural affiliation, and self-identification. As emphasized by Bhopal (2004), ethnicity reflects “a mix of cultural and other factors including language, ancestry, and self-identity,” and researchers are encouraged to classify ethnic groups using “one or a mix” of these indicators [58]. This conceptualization aligns with long-established Sámi health research traditions, which operationalize Sámi ethnicity using both linguistic and self-identification dimensions [59].

The combined use of language indicators and self-identification is particularly crucial in the Sámi context due to the historical effects of Norwegian assimilation policies. These policies contributed to the loss of Sámi language transmission and to shifts in ethnic self-identification across generations, resulting in situations where individuals may have Sámi ancestry but do not identify publicly or subjectively as Sámi. Findings from the North Norwegian Youth Study [46] illustrate this divergence clearly: fewer than half of adolescents with a Sámi ancestral background reported Sámi self-identification. Consequently, relying solely on either ancestry/language background or self-identification would systematically misclassify a substantial proportion of Sámi youth.

In line with this evidence, the present study classifies adolescents as Sámi if they meet either of the two criteria, or both. The first criterion is Sámi language use at home. Respondents will indicate spoken language (Sámi, Norwegian, Kven, Finnish, Swedish, or other), with multiple responses permitted, and participants will be classified as Sámi when Sámi is reported as a language they have learned or use at home. The second criterion is self-identification, defined by selecting Sámi—either alone or in combination with other categories—in response to the item “I regard myself as…,” which offers the same set of ethnic options and allows multiple responses.

Those who report no Sámi language affiliation and do not identify themselves as Sámi will be classified as non-Sámi. This group comprises primarily majority Norwegian adolescents and ethnocultural Kven youth, reflecting the population composition of Finnmark. A small number of adolescents with other non-Sámi backgrounds (e.g., immigrant or minority backgrounds not Norwegian or Kven) will be included in the non-Sámi category; these groups will not be examined separately due to limited sample size and will not be the focus of analyses.

To enhance transparency and acknowledge potential heterogeneity within the Sámi group, descriptive analyses will distinguish adolescents who meet only the language-use criterion, only the self-identification criterion, or both.

### Problematic internet use (PIU)

The Problematic Internet Use Questionnaire, Short Form (PIUQ-9), will be used to measure problematic internet use (PIU) [60, 61]. The PIUQ is a widely used and psychometrically robust instrument, making it a preferred choice in research assessing PIU [62, 63]. This validated instrument assesses three dimensions of problematic internet use: obsession, neglect, and control disorder, with three items per dimension [60]. The obsession dimension captures obsessive thinking about the internet, including daydreams, fantasies, and mental withdrawal symptoms when access is restricted. The neglect dimension reflects the disregard of essential needs and everyday activities, such as sleep, schoolwork, and in-person interactions. The control disorder dimension assesses difficulties in regulating internet use, including unsuccessful attempts to reduce or control online behavior. Each item is rated on a 5-point Likert scale (1 = "never" to 5 = "always"), resulting in a total score ranging from 9 to 45, with higher scores indicating greater levels of problematic internet use.

For descriptive prevalence reporting, a cut-off score of ≥ 22 will be pre-registered to classify participants as being "at risk" of problematic internet use. This threshold is based on proportional scaling from the established cut-off for the PIUQ-18 (≥ 41) [60]. Although this cut-off will be used to estimate prevalence, PIU will be treated as a continuous latent variable in all structural models to capture the full range of variability in participants’ responses. Both the PIUQ-9 and PIUQ-18 have demonstrated satisfactory reliability and validity across diverse cultural contexts [60, 62]. However, no universally established cut-off score exists for the PIUQ-9, and prior studies have highlighted the need to develop such thresholds to enhance its applicability [62, 64]. Given the variability in cut-off recommendations across study countries, the cut-off of ≥ 22 will be used as a descriptive benchmark rather than a diagnostic criterion.

To ensure contextual relevance to contemporary internet use—particularly considering changes in youth internet behavior such as widespread smartphone use—the PIUQ was adapted based on input from group interviews with Sámi and non-Sámi high school students. These revisions updated the original questionnaire, which was developed over a decade ago. Permission for adaptation to the Finnmark context was obtained from the original author.

### Positive internet use

In addition to assessing problematic internet use, the questionnaire includes items on positive internet use. Positive internet use is conceptualized as a conceptually and empirically informed construct derived from previous research on Indigenous youth. These items are informed by previous research showing that digital technologies can support social connection, emotional support, cultural engagement, and identity affirmation among Indigenous youth in Australia and Canada [22–24].

Community belonging and cultural connectedness have also been identified as protective factors associated with well-being and resilience among Indigenous adolescents [26]. Therefore, positive internet use in the present study is conceptualized as adolescents’ use of digital technologies to maintain social connections, seek social support and emotional connection, and engage with cultural identity. These domains were further refined through group interviews with Sámi and non-Sámi adolescents to ensure cultural relevance in the Finnmark context.

The pilot phase will be used primarily to assess item clarity, relevance, and feasibility rather than to establish the psychometric properties of the construct. The dimensionality, reliability, and construct validity of the positive internet use items will subsequently be evaluated in the full study sample prior to inclusion in structural analyses. If the items demonstrate a unidimensional structure, they will be modeled as indicators of a single latent construct. If a multidimensional structure emerges, positive internet use will be modeled as a higher-order latent construct.

### Social norms

All social-norm items used in this study are operationalized as *perceived norms*—that is, adolescents’ subjective perceptions of others’ behaviors (descriptive norms) and approvals or disapprovals (injunctive norms) within family, peer, and school contexts, rather than objective norms obtained directly from those referent groups.

Social norms related to internet use behaviors will be measured using items adapted from theories of social norms [32, 33] and previous research on adolescent risk-taking behaviors [34, 35, 37–39]. Descriptive norms measure how frequently participants believe their reference group engages in internet-related behaviors; reference groups are defined as the cultural and social networks that adolescents consider when making behavioral decisions [32, 33]. Injunctive norms will assess perceived approval or expectations from their reference group regarding internet use. Social norms items were developed through group interviews with Sámi and non-Sámi high school students to ensure cultural relevance and alignment with current internet use. The study focuses on three key reference groups influencing adolescents use of internet and mobile phone: (1) Family—parents and older household members; (2) Peers—both in-person and online friends; and (3) School — teachers and rules regarding internet and mobile phone use. Assessing norms across these domains provides a comprehensive view of the social influences associated with adolescent internet behaviors.

### Ethnic identity and ethnic-related victimization

Ethnic identity will be assessed using the revised version of the Multigroup Ethnic Identity Measure (MEIM) [65], which captures two core dimensions: ethnic identity search (cognitive exploration) and affirmation, belonging, and commitment (affective attachment). This instrument has been widely applied in diverse cultural settings [66, 67], with the Norwegian versions applied in NNYS [51] and NAAHS [31]. Items assessing ethnic-related victimization, such as experiences of discrimination, will be adapted from NAAHS [31, 68]. Permission to use questionnaire items from NNYS and NAAHS has been obtained from the original authors.

### Control variables

To reduce potential confounding, covariates were selected a priori based on prior research on adolescent internet use [43, 46, 69]. Individual-level control variables include gender, grade level (VG1–VG3; equivalent to 11th–13th), living arrangement (with parents vs. dormitory), depressive symptoms, alcohol use, smoking/nicotine use, and ethnic community context.

Depressive symptoms are measured using five items assessing emotional distress during the past week, rated on a four-point scale and treated as a continuous measure. Alcohol use is assessed by frequency of consumption (*never* to *weekly*) and intoxication during the past 12 months (*never* to *>10 times*). Smoking/nicotine use is measured by current smoking frequency (*never smoked* to *weekly or more*). All items are drawn from the National Norwegian Youth Survey (Ungdata) and included for statistical adjustment in the structural models.

Ethnic community context is included as a school-level control variable, operationalized by whether the school is located in a municipality included in the Sámi Language Act administrative area [70] (used as a proxy for higher Sámi density). Schools located in Kautokeino, Karasjok, Lakselv (Porsanger), and Tana are included in this administrative area and are therefore classified as higher Sámi density, whereas schools in all other municipalities are classified as lower Sámi density. This variable is entered as an exogenous covariate with direct paths to the outcome variables and is not used for subgroup or multigroup analyses due to sample size considerations.

### WP2: Data collection

Data will be collected from all ten high schools in Finnmark County, located in Alta, Hammerfest, Honningsvåg, Karasjok, Kautokeino, Kirkenes, Lakselv, Tana Bru, Vadsø, and Vardø. Sampling design and expected numbers are described above.

Recruitment of participants will be conducted in the third to fourth quarters of 2026. The data collection period is scheduled to commence in the first quarter of 2027 and will continue for one year.

The survey will be conducted anonymously and voluntarily in a classroom setting during school hours, with a research team member present providing instructions, answer questions, and ensure compliance with ethical guidelines. The primary mode of administration will be digital; however, paper-based questionnaires will be available for students who prefer a non-digital format.

Before data collection begins, school administrators and teachers will be informed about the study’s objectives, procedures, and ethical considerations to ensure smooth coordination. Students will receive an information sheet outlining the study’s purpose, data confidentiality, and their right to withdraw at any time without consequences. No time limit will be imposed to answer the questions, allowing participants to complete the questionnaire at their own pace. Following data collection, all responses will be securely stored in accordance with institutional data protection requirements.

### WP3: Data analysis

#### Data preparation

Prior to analysis, the collected data will undergo a thorough cleaning and preparation process to ensure accuracy and reliability. This process includes examining the distribution of values, identifying potential collinearity, and addressing missing values, outliers, and inconsistencies.

The extent and pattern of missingness will be examined across all variables included in the PLS-SEM measurement and structural models. Following published PLS-SEM recommendations, respondents with more than 15% missing data across all model-relevant indicators will be excluded prior to imputation. This threshold aligns with guidance that variables or cases with excessive missingness should be removed to maintain data quality. The percentage of missingness per variable and for the full dataset is transparently reported, as recommended [71].

For remaining missing values, multiple imputation by chained equations (MICE) will be used under the assumption of missing at random (MAR), generating m = 30 imputed datasets. This approach is consistent with best practices, which recommend multiple imputation (or maximum likelihood-based methods) over deletion when missingness exceeds minimal levels (∼5%) and strongly advocate for imputation when missingness surpasses ∼10% to preserve statistical power and reduce bias. The imputation model included all variables used in the analysis as well as auxiliary predictors of missingness to improve the accuracy of imputations [71–73].

Because the study involves multigroup PLS-SEM (PLS-MGA) to compare Sámi and non-Sámi adolescents, imputations will be conducted separately within each group. This ensures that group-specific distributions, means, variances, and covariances are preserved, which is critical for valid multigroup comparisons and measurement invariance testing (MICOM). This procedure follows PLS-SEM guidance on carefully handling grouping variables during preprocessing to avoid introducing biased or artifactual differences [71, 72].

Outliers will be identified and treated following PLS-SEM data quality recommendations.

Suspicious response patterns (e.g., straight-lining, alternating extremes) will be screened using descriptive statistics and visual inspection, and any removals will be documented. Univariate outliers will be flagged using boxplots (values beyond three times the interquartile range [IQR]), while multivariate outliers will be assessed using Mahalanobis distance and leverage diagnostics appropriate for SEM preprocessing. Clearly erroneous outliers, such as data entry artifacts, will be removed.

However, extreme values that could be meaningfully justified will be retained, consistent with recommended practices. The final analytic sample size (N) is reported [71, 72].

To ensure consistency across numerical variables, all data will be standardized to a zero mean and a standard deviation of 1 prior to analysis.

### Descriptive statistics

Descriptive statistics (e.g., means, standard deviations, frequencies) will be calculated for all key variables, including PIU (total PIUQ-9 score), positive internet use, and demographic characteristics and control variables. Prevalence estimates for PIU will be reported using the pre-registered cut-off score of ≥ 22 on the PIUQ-9 to classify participants as being "at risk" of problematic internet use. Group comparisons (e.g., Sámi vs. non-Sámi adolescents) will be conducted using non-parametric tests, such as the Kruskal-Wallis H test, to account for potential differences in sample sizes and distributions.

### Partial Least Squares Structural Equation Modeling (PLS-SEM)

PLS-SEM analysis will be performed to investigate the associations between exogenous and endogenous variables. PIU will be modeled as a continuous latent construct with three reflective dimensions (obsession, neglect, and control disorder), while positive internet use will be modeled as a separate latent construct indicated by items reflecting social connections, emotional support, and cultural engagement.

As the first step of the PLS-SEM analysis, model evaluation will be conducted for both the measurement and structural models. Assessing the measurement models included evaluating factor loadings, reliability and convergent validity. Factor loadings indicate the variance explained by a measured variable on a specific factor, with factor loadings of 0.5 or higher considered acceptable [55]. Internal consistency reliability will be evaluated using composite reliability (CR) for each latent variable. Unlike Cronbach’s alpha, which assumes equal indicator loadings, CR offers more flexibility and is thus more suitable for PLS-SEM. [55]. Convergent validity will be evaluated using the average variance extracted (AVE), with values of 0.4 or higher indicating adequate convergence [55]. Once the measurement model is validated, the structural model will be analyzed to test the study’s hypotheses.

Bootstrapping with 5,000 resamples will be used to generate confidence intervals and determine statistical significance (p < 0.05) [55]. Multicollinearity among predictor constructs will be assessed using the variance inflation factor (VIF), with values above 5.0 indicating potential collinearity [55].

### PLS Multi-Group Analysis (PLS-MGA)

After evaluating the structural model, we will examine whether the relationships among the study variables differ between Sámi and non-Sámi adolescents through Partial Least Squares Multi-Group Analysis (PLS-MGA). Because meaningful multigroup comparisons require that constructs are measured equivalently across groups, the analysis begins with an assessment of measurement invariance using the Measurement Invariance of Composite Models (MICOM) procedure [74, 75]. In this step, we first ensure that both groups share the same basic measurement configuration, including identical indicators, model specifications, and data treatment procedures. We then evaluate whether the composites are formed in a comparable way across groups by testing the equivalence of their indicator weights through a permutation-based approach. Finally, we assess whether composite means and variances differ statistically across groups, which allows us to determine the level of invariance established before proceeding to the substantive group comparisons.

The primary preregistered multigroup analysis uses a combined definition of Sámi identity, classifying adolescents as Sámi when they meet either the language-use or self-identification criterion, or both. To enhance transparency and assess robustness, descriptive analyses will distinguish between adolescents who meet only the language-use criterion, only the self-identification criterion, or both. In addition, planned sensitivity analyses will restrict the Sámi group to adolescents who meet both criteria.

Following establishment of measurement invariance, multigroup structural comparisons will evaluate whether associations among social norms, ethnic identity, ethnic victimization, and problematic or positive internet use differ between groups. These comparisons draw on established PLS-MGA procedures [75, 76], including permutation-based assessments of whether observed between-group differences exceed what would be expected by random reassignment, bootstrap-based evaluations of group-specific sampling distributions, and complementary parametric tests that examine differences under assumptions of equal and unequal variances. Together, these procedures allow for a comprehensive assessment of potential group-specific differences in both measurement and structural relations.

By integrating MICOM with PLS-MGA, the analysis provides a rigorous basis for determining whether associations among social norms, ethnic identity, ethnic victimization, and problematic or positive internet use vary between Sámi and non-Sámi adolescents.

To reduce potential confounding, the primary PLS-SEM specification will include the following exogenous covariates with direct paths to PIU: gender, grade level (VG1–VG3), living arrangement (with parents vs. dormitory), depressive thoughts, alcohol use, smoking, and Sámi community context.

### WP4: Dissemination

The study’s findings will be disseminated through academic, institutional, school, and community platforms, including locally tailored summaries. Three to four peer-reviewed articles are planned: the first will present the methodological and theoretical framework; the second will report descriptive findings and group comparisons; the third will test hypotheses related to social norms, ethnic identity, and victimization as predictors and moderators of PIU using PLS-MGA. Additional articles may focus on subgroup analyses or the full-sample structural model. Findings will also be presented at national and international conferences.

### Project timeline

The project is scheduled to run from 2026 to Q1 2029 (a full three-year workload) and structured by quarterly intervals, as shown in Table 1. Key work packages include questionnaire development and piloting, data collection and analysis, and dissemination of results. Activities such as instrument refinement, pretesting, and translation are scheduled early in the project, followed by structured phases of data collection, analysis, and ongoing dissemination through academic channels.

**Table 1:**
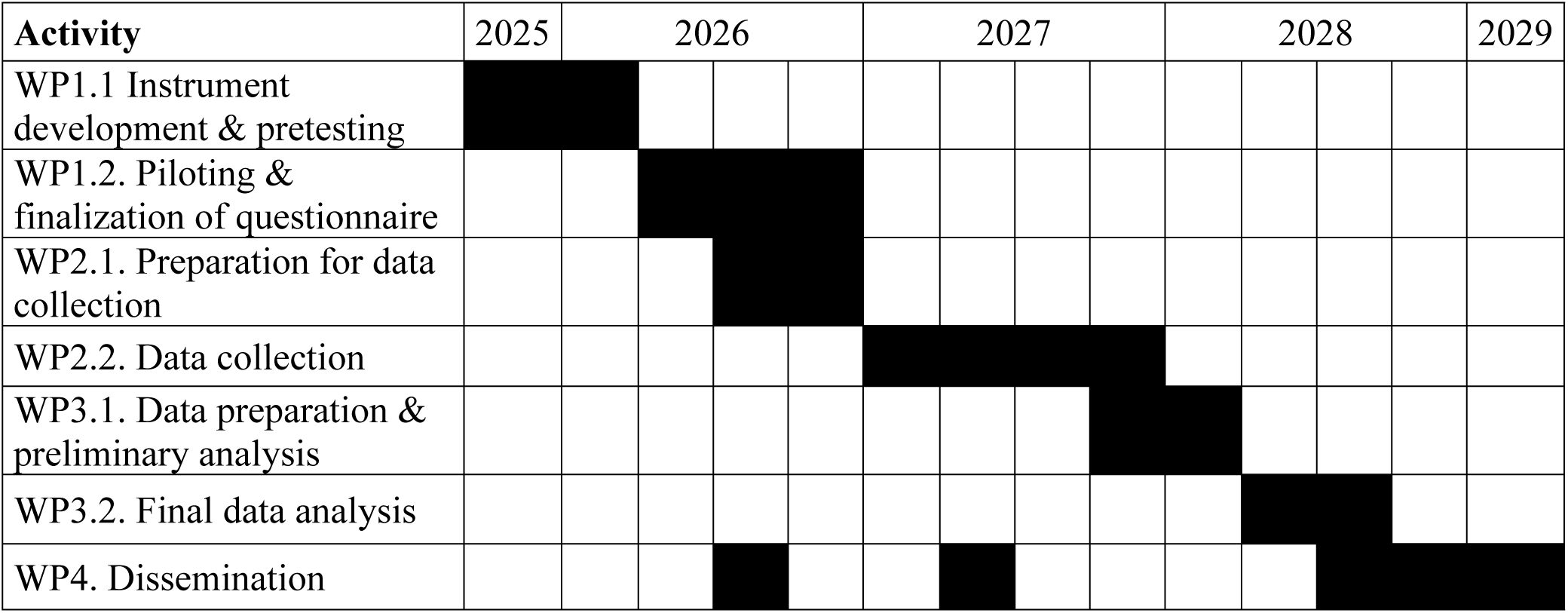
Gantt chart (quarterly plan)

### Ethics and data protection

Given the sensitivity of topics such as ethnic background, language use, and experiences of discrimination, this study adheres to national and international ethical guidelines for research involving Indigenous populations. Emphasis is placed on respect, cultural sensitivity, and informed, voluntary participation. Ethical oversight was provided by the Data Protection Officer at Finnmark Hospital Trust, serving as the institutional equivalent of an ethics review board (IRB) (Ref. Nr. 0234). Collective Sámi consent was obtained from the Expert Ethics Committee for Sámi Health Research.

The project processes special categories of personal data—specifically, ethnicity and language background—under Article 9(2)(j) of the General Data Protection Regulation (GDPR), which permits processing for scientific research in the public interest with appropriate safeguards in line with Article 89(1). In Sámi contexts, language may indirectly reflect ethnic origin, while self-identification as Sámi is treated as explicit ethnic data. These variables are essential for studying ethnic identity.

All participants will be over 16 years of age and will provide their own informed consent in accordance with Norwegian regulations. Consent will be obtained either in written (paper) or digital form. Participants will indicate their agreement by checking the consent declaration form without providing names or other personal identifiers, as the survey will be conducted anonymously. Study information and consent forms will be made available in both Norwegian and Northern Sámi. No minors will be included in the study, and therefore parental or guardian consent is not required.

Data are collected without direct identifiers and stored in pseudonymized form to allow necessary linkage while minimizing privacy risk. Digital responses are captured via Research Electronic Data Capture (REDCap), a secure, web-based platform with encrypted storage and access controls. Data are stored on encrypted servers at Finnmark Hospital Trust with multi-factor authentication and audit logging. Only authorized personnel have access. Upon project completion, all data will be anonymized.

Sensitive data are further processed within secure environments such as the Services for Sensitive Data (TSD), in line with GDPR. Non-sensitive communications are conducted via Microsoft 365 under institutional agreements. Data will not be used for commercial purposes, and no biological material is collected. Any future research use will require separate ethical approval and adhere to GDPR and institutional protocols.

### Risk-benefit assessment

The study involves no physical intervention and is fully pseudonymized. However, topics such as ethnic identity, discrimination, and problematic internet use may evoke emotional discomfort, particularly among Sámi adolescents. To mitigate this, participation is voluntary, questions may be skipped, and support staff will be available during school-based data collection. All data are securely stored and accessible only to authorized researchers, as described above. The study follows ethical principles for Indigenous research, emphasizing relevance, respect, and voluntary involvement.

The potential societal benefits—particularly improved understanding of internet use, social norms, and ethnic identity—outweigh the minimal risks. Findings may inform culturally adapted support programs in schools and health services, promoting inclusion and well-being.

### User participation

Representatives from high school students—both Sámi and non-Sámi—as well as school administrators will be actively involved throughout multiple phases of the research process. The first step involved adapting existing instruments, such as the PIUQ, and developing contextually appropriate measures of social norms, with students from both groups interviewed to provide feedback. A draft questionnaire was then developed. Pretesting will follow, during which participants will provide feedback to refine wording and structure to improve the overall quality of the questionnaire.

Prior to data collection, informational meetings will be held with school staff and students to explain the study and address any questions. A data collection plan will be developed in collaboration with participating schools to support recruitment and student engagement.

### Competence, infrastructure, and resources

The study is hosted by the Sámi National Competence Centre for Mental Health and Substance Use (SANKS) and Finnmark Hospital Trust, both of which provide an established research infrastructure. This includes access to TSD, the REDCap platform for data collection, and administrative support for ethics applications, and data management.

The project team comprises interdisciplinary members with clinical and research expertise in psychology, psychiatry, addiction medicine, and Sámi youth mental health. Team members have strong competence in Sámi topics and cultural contexts, ensuring that both the Sámi dimension and the topic of adolescent internet use are meaningfully integrated throughout the research process. The core research team includes psychiatrists and a licensed specialist in clinical psychology with extensive clinical and research experience related to Sámi child and youth mental health and substance use. Core staff are responsible for study design, cultural adaptation of instruments, and coordination with participating schools. An advisory group contributes additional expertise in adolescent mental health, online behavior, and regional public health. An authorized Sámi translation service provides professional translation between Norwegian and Northern Sámi.

## Discussion

This study presents an investigation into Problematic Internet Use (PIU) among Sámi and non-Sámi adolescents in Finnmark, Norway. Drawing on Social Norm Theory and Ethnic Identity Theory, it examines how digital behaviors are shaped by normative expectations within family, peer, and school contexts, and how ethnic identity and experiences of victimization function as risk or resilience factors.

A cross-sectional design and total population sampling ensure broad representation from adolescents in a geographically sparsely populated, multiethnic region. Partial Least Squares Structural Equation Modeling (PLS-SEM) supports the analysis of complex relationships among latent variables and allows for group comparisons. A bilingual, culturally adapted questionnaire increases the relevance and accessibility of the study for Sámi participants. Dissemination will take place through academic, institutional, school, and community platforms, with tailored outputs for Sámi stakeholders.

The study adheres to established ethical and data protection standards, including compliance with GDPR, Indigenous research ethics, and institutional safeguards. Approvals from general and Sámi-specific ethics committees provide oversight and ensure cultural and legal integrity.

By addressing a gap in the literature on PIU in Indigenous youth, the study contributes knowledge on modifiable social norms and the protective role of ethnic identity. These findings offer a basis for culturally informed strategies in school health services, mental health care, and community-based prevention. The study has relevance for both regional and broader efforts to promote digital well-being among adolescents in Arctic, rural, and Indigenous contexts.

Any major amendments to the study protocol (e.g., revisions to the questionnaire, analysis plan, or participant recruitment) will be submitted to the appropriate ethics committees (both general and Sámi-specific) for review and approval. In the case of unforeseen circumstances requiring significant deviation from the protocol or early termination of the study, documentation will be provided to the funding body and relevant ethics boards. Participants and stakeholders will be informed of any changes that may affect their involvement.

Several limitations must be acknowledged. First, the cross-sectional design restricts the ability to establish causal relationships. While associations among social norms, ethnic identity, victimization, and PIU will be analyzed, the temporal sequence of these factors remains unclear. Longitudinal research is needed to determine directionality and causality.

Second, the study relies on self-reported data, which are susceptible to biases such as recall inaccuracies and social desirability. Although anonymity is ensured to reduce such effects, the risk of response bias remains. Furthermore, measures of social norms are based on participants’ perceptions of others’ attitudes and behaviors, which may not reflect actual group norms. In addition, all social-norm measures capture perceived norms—adolescents’ subjective interpretations of others’ behaviors and attitudes—which may differ from objective norms reported directly by parents, peers, or teachers.

These discrepancies may influence the strength or direction of observed associations and should be considered when interpreting results. Although the questionnaire will be translated and culturally adapted, the number of participants completing the Northern Sámi version may be insufficient for comprehensive testing of measurement invariance across language versions. Therefore, potential language-related measurement differences cannot be fully excluded.

Third, while the sampling strategy targets the entire population of high school students in Finnmark, participation is voluntary. If certain subgroups (e.g., those with low school attachment or reluctance to disclose ethnic identity) are underrepresented, the generalizability of findings may be limited. In addition, because the study includes only adolescents currently enrolled in high school and present on the day of data collection, it does not capture youth who were absent (e.g., truancy, irregular attendance, or sleep-related lateness possibly linked to PIU) or those who have already dropped out—a group that may have higher levels of problematic internet use. This is particularly relevant in Finnmark, where high school completion rates are lower than the national average (approximately 75% compared with about 86% in Oslo [77]), despite recent improvements. Prior research indicates that individuals with more severe PIU are more likely to be out of education or employment [61], suggesting that the prevalence of PIU in our sample may underestimate the true burden among all adolescents in Finnmark. Furthermore, although a PIUQ-9 cut-off score of ≥22 will be used for descriptive prevalence estimates, this threshold is derived from proportional scaling of the PIUQ-18 and has limited psychometric validation. Consequently, prevalence estimates should be interpreted with caution.

The classification of adolescents as Sámi or non-Sámi was used as an analytical tool rather than as a comprehensive measure of ethnic identity. As such, this operationalization may oversimplify the fluid and situational nature of ethnic identity, particularly in contexts shaped by historical assimilation. Findings related to ethnic differences should therefore be interpreted with this limitation in mind.

Finally, the use of PLS-SEM—though appropriate for complex modeling and small sample sizes—entails multiple statistical tests, which increases the risk of type I error. While bootstrapping and robustness checks will be applied, the possibility that some observed associations may represent false-positive findings arising from multiple statistical testing should be considered. Moreover, although PLS-SEM allows for modeling complex latent structures and multiple relationships simultaneously, it does not resolve the temporal ambiguity inherent in cross-sectional data; therefore, all modeled associations should be interpreted as correlational rather than causal.

In conclusion, this study offers a culturally grounded investigation into Problematic Internet Use among Sámi and non-Sámi adolescents in a multiethnic Arctic context, with a focus on social norms, ethnic identity, and victimization.

## Authors’ contributions

SH had overall responsibility for developing the research protocol, including conceptualization, securing funding, designing the methodology, coordinating the project, and writing the manuscript. SM contributed to the study’s conceptual and methodological development and provided input on the manuscript. ARS contributed to the development of the conceptual framework, curated relevant research resources, and provided input on the manuscript. CJ provided general oversight and support across phases (planning, conceptualization, writing) and offered resources. All authors reviewed and approved the final version of the manuscript.

## Acknowledgements

We thank the young people who participated in the participatory meetings for their valuable contributions to the development of the questionnaire. We further acknowledge Ronald Isaksen, Administrative Coordinator at the Sámi Norwegian National Advisory Unit for Mental Health and Substance Use (SANKS), for his role in organizing and facilitating meetings with youth and schools. Finally, we appreciate the Research Department at Finnmark Hospital Trust for providing a supportive and enabling research environment.

## Declaration of generative AI and AI-assisted technologies in the writing process

During the preparation of this work, the author used Copilot to improve the readability and language of the manuscript, as well as to check grammar.

## Data availability

Upon study completion, de-identified survey data and the corresponding codebook will be deposited in the Norwegian Research Information Repository (Nasjonalt vitenarkiv, NVA), the national open-access archive for research outputs operated by Sikt (Norwegian Agency for Shared Services in Education and Research). Data will be shared in accordance with PLOS ONE’s data availability policy.

